# Facemask usage during the COVID-19 pandemic among people with primary ciliary dyskinesia: a participatory project

**DOI:** 10.1101/2021.04.13.21255393

**Authors:** Eva SL Pedersen, Eugenie NR Collaud, Rebeca Mozun, Katie Dexter, Catherine Kruljac, Hansruedi Silberschmidt, COVID-PCD patient advisory group, Jane S Lucas, Myrofora Goutaki, Claudia E Kuehni

## Abstract

**Background:** Facemasks help prevent transmission of SARS-CoV-2 and are particularly important for people with chronic respiratory disease such as primary ciliary dyskinesia (PCD). We studied the usage of facemasks and its consequences among people with PCD in an international context.

**Methods:** We used data from COVID-PCD, an international observational cohort study which collects longitudinal data from people with PCD during the COVID-19 pandemic via weekly online questionnaires. An online questionnaire investigating use of facemasks was posted in October 2020. It asked about frequency of facemask usage in different public places, problems experienced with use of facemasks, affordability of masks, and beliefs regarding their effectiveness.

**Results:** 282 participants (50%) completed the questionnaire. Participants came from 27 different countries; median age was 32 years (interquartile range 17-48), and 63% were female. Almost all wore a facemask whenever they left their house. In addition, many avoided public places altogether. For example, 159 (57%) did not use public transport at all, 108 (39%) always wore a facemask in public transport, 4 (1%) sometimes, and 7 (3%) never. 81% agreed with the statement that facemasks protect the person who wears the mask, and 91% agreed that facemasks protect others. One third reported that it was uncomfortable to wear a mask because of their runny nose, 31% because of cough, and 22% because of difficulty breathing. Participants less often wore facemasks in public when there was no national requirement. Only two persons had a personal exemption from wearing a facemask prescribed by a physician.

**Conclusions:** This international study found that people with PCD carefully shield themselves, and most wear facemasks everywhere in public. People who did not wear facemasks in public came from countries without a national facemask requirement. National policies mandating facemask use in public are important for universal use to protect high-risk populations from SARS-CoV-2 infections.

## Background

Facemasks help prevent transmission of SARS-CoV-2 between people [1-4] and have become a key preventive measure in the COVID-19 pandemic [5, 6]. Many governments around the world implemented mandatory facemask use in public spaces throughout 2020 [7]. Facemasks are especially important for people who are at high risk of severe COVID-19, such as elderly people or those with chronic diseases [8-10]. However, many people have reported discomfort resulting from the use of facemasks such as difficulty breathing, pain around the ears, headaches, or skin problems [11, 12]. High-risk populations such as people with a chronic respiratory disease may experience additional problems because of respiratory symptoms such as chronic cough and rhinitis. There are no studies reporting on the frequency of facemask usage or on problems encountered by people with a chronic respiratory disease.

Primary ciliary dyskinesia (PCD) is a rare genetic multi-system disorder, where dysfunctional cilia lead to impaired mucociliary clearance, laterality defects, and other health problems [13-17]. Most people with PCD have recurrent upper and lower airway disease resulting in chronic respiratory symptoms such as constantly runny nose and chronic cough with sputum production [13, 18]. Lung function in people with PCD is often reduced and can lead to oxygen requirement [19-22]. Hearing impairment is common because of chronic otitis media [16, 23]. At the start of the pandemic, people with PCD were considered at high risk of a severe disease course if infected with SARS-CoV-2 and were therefore recommended to ensure good shielding, including using facemasks. People with PCD might be particularly burdened by facemasks due to chronic wet cough, constantly runny nose, hearing problems, or psychological reasons such as fear of stigmatization. We aimed to understand the usage of facemasks and its consequences among people with PCD in an international context. Specifically, we assessed how often and which type of facemasks people with PCD wear, studied beliefs related to facemasks, described problems reported in relation with their use, and described characteristics of participants who did not wear a facemask in public.

## Material and methods

### Study design and inclusion criteria

We used data from COVID-PCD, an international observational cohort study that uses anonymous online questionnaires to collect information directly from people with PCD during the COVID-19 pandemic (clinicaltrials.gov: NCT04602481). COVID-PCD is a participatory research project where people with PCD have an active role in all stages of research from the design of the study, its content, the piloting, and communication of results. Details about the study methods have been published [24, 25]. In short, the COVID-PCD study includes persons of any age from anywhere in the world with a confirmed or suspected diagnosis of PCD. The study is designed for three age groups; children below 14 years, adolescents between 14 and 17 years, and adults aged 18 years or more. For children, the questionnaires are addressed to the parents, but the child is encouraged to help complete the questionnaires. Adolescents and adults complete the questionnaires themselves. The study is available in English, German, Spanish, Italian, and French. Recruitment started on May 31, 2020. The Cantonal Ethics Committee of Bern approved the study (Study ID: 2020-00830). Informed consent to participate is provided online at the time of registration into the study. This article follows the STROBE reporting recommendations [26].

### Study procedures

The COVID-PCD study is conducted online. Participants are invited by PCD support groups who contact and inform people living with PCD through social media and email networks and encourage them to take part. The website (www.covid19pcd.ispm.ch) includes detailed information about the study and allows participants to register and consent via a link that leads them directly to the study database. Participants first complete a baseline questionnaire with questions on their disease, their usual symptoms, and SARS-CoV-2 infections experienced prior to joining the study. Thereafter they receive weekly follow-up questionnaires with questions on incident SARS-CoV-2 infections, current symptoms, social contact behaviour, and physical activity. Intermittently, questionnaires focus on special topics. This paper presents data from a special questionnaire on facemasks that was sent to participants on October 10^th^ 2020. Participating PCD support groups were strongly involved in the development of the questionnaire. Participants received up to two reminders if they did not respond to the first questionnaire. They answered questions online, and data were saved in a Research Electronic Data Capture (REDCap) database, developed at Vanderbilt University (www.project-redcap.org) [27], which is securely hosted by the Swiss medical registries and data linkage centre (SwissRDL) at the University of Bern, Switzerland.

### Information about mask use

The questionnaire asked whether participants used facemasks in public and if so, in which places, and whether they were exempt from wearing a face mask because of their disease (**additional file 1**). We also asked about problems people with PCD experienced when wearing facemasks and whether they needed to remove their mask because of these problems. We asked about their beliefs regarding effectiveness of facemasks, whether participants experienced communication difficulties, and whether costs for facemasks represented a financial burden to them. In the questionnaire for children, we asked if the child was too young to wear a facemask.

We collected information about facemask requirements in different countries through masks4all (https://masks4all.org/) [7].

### Statistical analyses

We described demographics of the participants, frequency of face mask use, and problems related to wearing a facemask using number and proportion for categorical variables and mean and standard deviation (SD) or median and interquartile range for continuous variables. We studied determinants of not wearing a mask in public by dividing participants into 3 categories, 1) always wear facemask, 2) does not wear facemask in one place, 3) does not wear facemask in two or more public places. These three categories were constructed from reported facemask usage in the following public places: grocery stores, clothes stores, restaurants, cinemas, hairdressers, physiotherapists, post-/bank offices. We selected these places because facemasks were compulsory in those places in most countries at the time of the study. We then compared characteristics of people always wearing face masks in public with people not wearing facemasks in one or more public places using chi-square and Fisher’s exact. We reported variables only for persons with valid information. Our dataset had few missing values (less than 1% in single variables) except for questions asking about who pays for masks (10% missing answers) and affordability of masks (15% missing answers). We used STATA version 14 for statistical analysis.

## Results

297 of the 572 people (53%) who participated in the COVID-PCD study in October 2020 returned the special questionnaire. We excluded 15 children because their parents reported them too young to wear a facemask regularly, and we finally included 282 people with PCD in this analysis. Median age was 32 years (age range: 3-85 years, interquartile range 17 to 48) (**table 1**). Participants who completed the facemask questionnaire were older than those who did not complete it (median age of non-responders: 22 years (interquartile range 8-37) (**additional file 2**). Study participants came from 27 different countries, with largest numbers from the UK (n=62; 22%), Germany (n=57; 20%), USA (n=45; 16%), and Switzerland (n=21; 7%). Facemasks were mandatory in some or all public places in 24 out of 27 countries at the time of the survey (**table 2**). Most participants used more than one type of facemask. Most common were reusable facemasks without exchangeable filters (n=136, 48%) followed by certified single use facemasks (n=128, 45%), and single use non-certified masks (n=56, 20%), filtering face piece (FFP, FFP2, or FFP3) (n=57, 20%), and fabric masks with exchangeable filters (n=64, 23%) (**table 1**). Few participants reported to have an exemption for wearing a facemask in public due to PCD; two participants (1%) had a personal facemask exemption prescribed by their physician and 32 (11%) reported that in their region people with PCD were exempt from wearing a facemask because of their chronic disease.

**Table 1:**
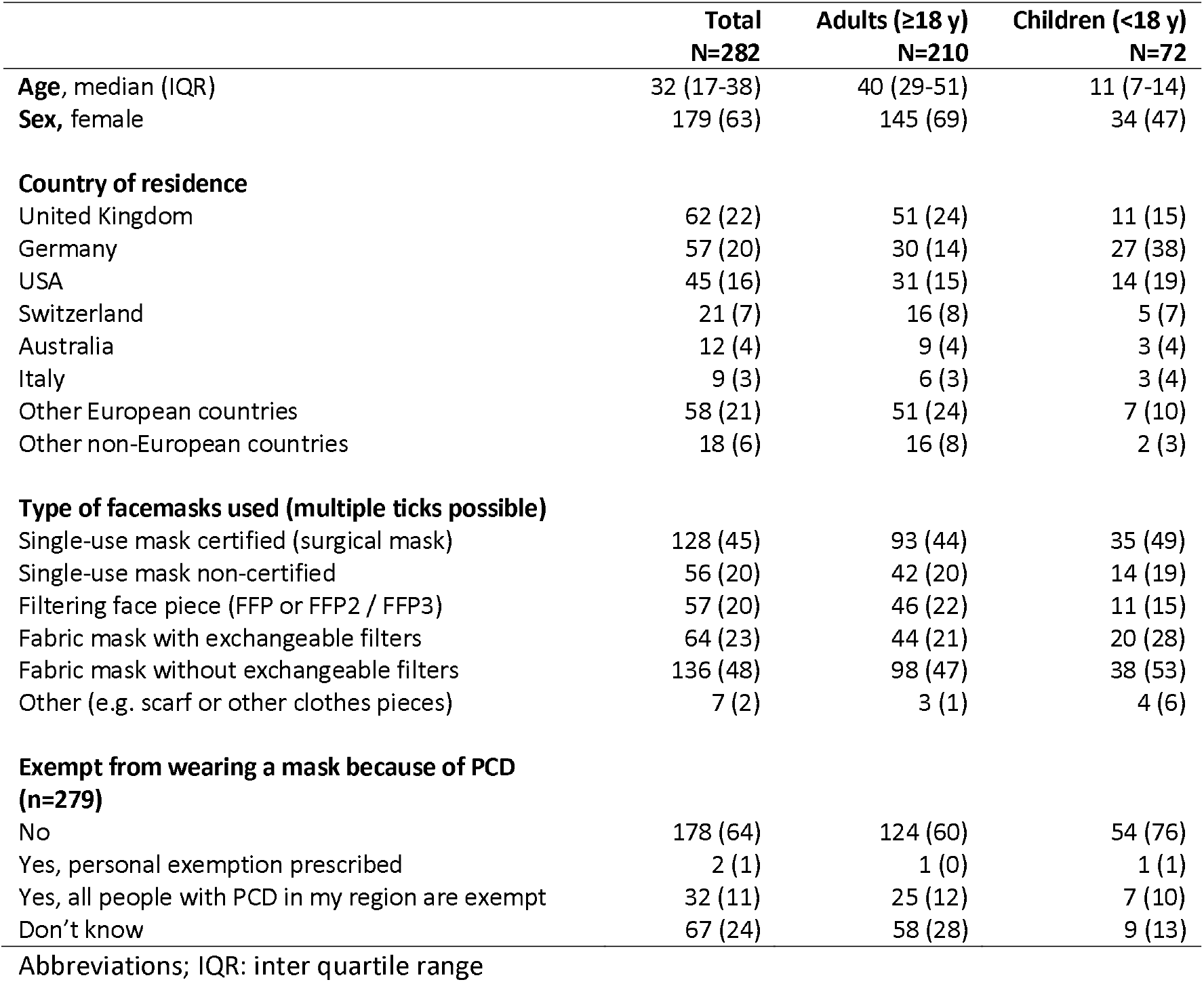
Demographic information, type of facemasks used, and exemption from wearing facemasks in total study population of people with PCD (n=282) and divided in adults (aged 18 years or older) and children (aged 17 years or younger).

**Table 2:** Number of study participants from different countries, number of people reporting that people with PCD are exempt from wearing facemasks in their region, and facemask requirements (status of 30.10.2020) and date of full country requirement in countries represented by study the participants.

| Country | No. of study participants<br>(total N=282) | People with PCD<br>are exempt from<br>wearing mask# | Mask required?* | Date of full country<br>requirement (yyyy-mm-dd)* |
| --- | --- | --- | --- | --- |
| Australia | 12 | 3 | Parts of country | - |
| Austria | 3 | 0 | Full country | 2020-03-30 |
| Belgium | 2 | 0 | Full country | 2020-05-06 |
| Brazil | 2 | 0 | Parts of country | - |
| Canada | 10 | 1 | Parts of country | - |
| Cyprus | 2 | 0 | Parts of country | - |
| Denmark | 6 | 3 | Full country | 2020-08-20 |
| France | 3 | 0 | Full country | 2020-05-11 |
| Georgia | 1 | 0 | Full country | 2020-04-20 |
| Germany | 57 | 0 | Full country | 2020-04-27 |
| Greece | 1 | 0 | Full country | 2020-04-27 |
| Hungary | 1 | 0 | No, but recommended | Mandatory from 2020-11-11 |
| Hong Kong | 1 | 0 | No, but recommended |  |
| Ireland | 4 | 1 | Full country | 2020-07-16 |
| Israel | 3 | 0 | Full country | 2020-04-01 |
| Italy | 9 | 1 | Full country | 2020-05-04 |
| Netherlands | 12 | 1 | Full country | 2020-06-01 |
| Norway | 6 | 1 | No |  |
| Poland | 4 | 1 | Full country | 2020-04-16 |
| Portugal | 1 | 0 | Full country | 2020-05-04 |
| South Africa | 1 | 0 | Full country | 2020-05-01 |
| Spain | 4 | 0 | Full country | 2020-05-02 |
| Sweden | 5 | 1 | No |  |
| Switzerland | 21 | 0 | Full country | 2020-04-03 |
| Turkey | 1 | 0 | Full country | 2020-04-03 |
| United Kingdom | 65 | 15 | Full country | 2020-06-15 |
| USA | 45 | 2 | Parts of country | - |

#reported by participants in study questionnaire \*Data source: https://masks4all.co/what-countries-require-masks-in-public/

Almost all participants wore a face mask whenever they left their house, but they also avoided many places with a high risk of transmission (**figure 1**). Taking public transport as example; 159 (57%) reported that they never used public transport, 108 (39%) reported that they always wore facemasks in public transport, 4 (1%) reported sometimes, and 7 (3%) reported never. The place visited by most participants was grocery stores where only 46 (16%) reported not going ever, 220 (79%) reported to always wear a mask, and 12 (4%) reported to never wear a mask. The place that was visited least often was fitness studios where 191 (78%) reported not to go ever, 31 (13%) reported always wearing a mask, and 18 (7%) reported never wearing a mask (**figure 1**). There was little difference between children and adults except that more children went to school and physiotherapy than adults while adults more often went to the bank or post office.

**Figure 1:**
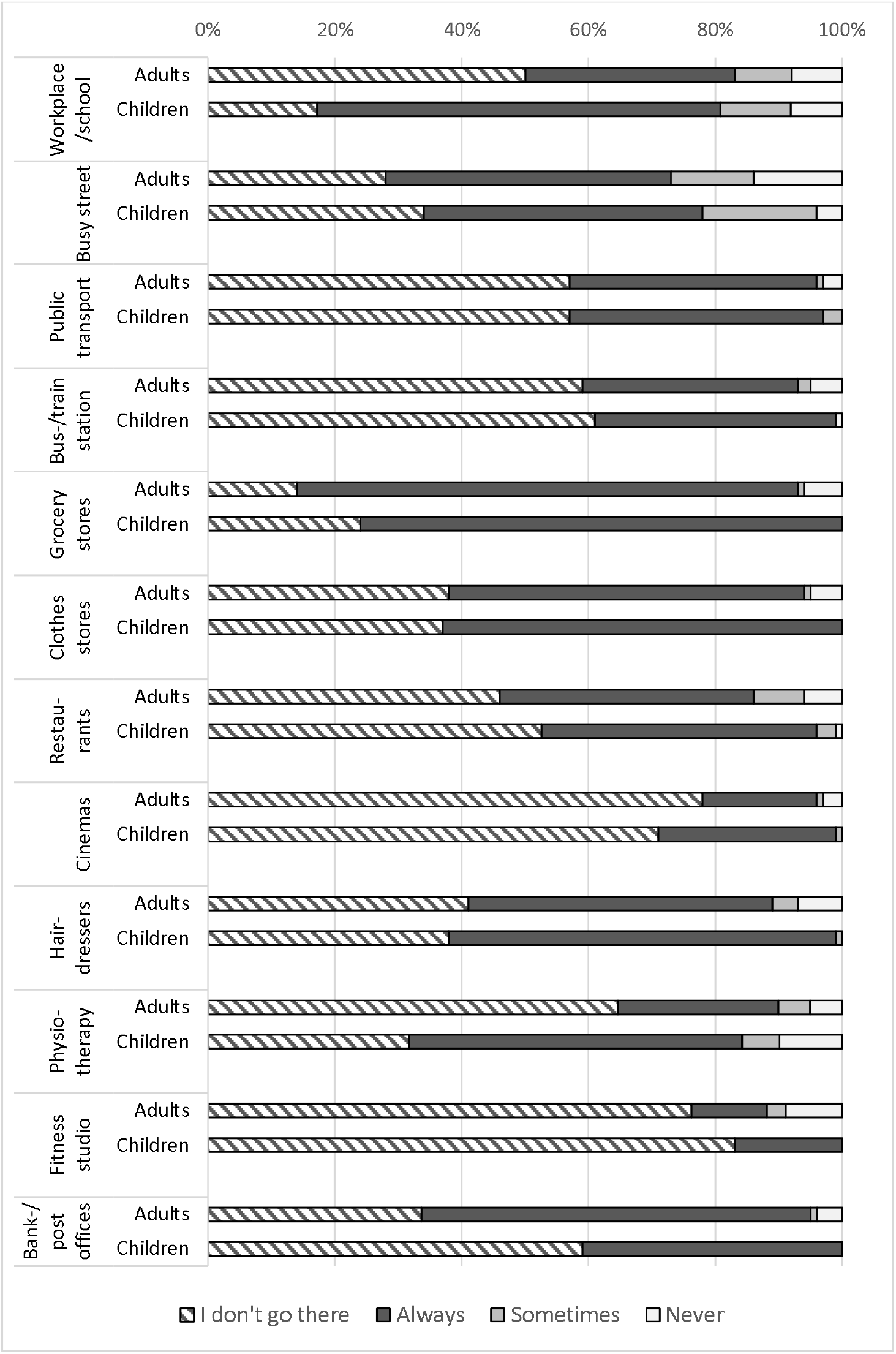
Frequency of people with PCD wearing a facemask at different places among adults aged 18 or older and among children and adolescents aged 17 years or below (self-reported, October 2020; N=282)

Most participants supported the view that face masks are effective in reducing transmission of SARS-CoV-2 with 227 (81%) agreeing that masks protect the person who wears the mask, and 257 (91%) agreeing that masks protect others from getting infected (**table 3**). Several reasons made it uncomfortable for people with PCD to wear a facemask. A third of participants (n=94) reported that it was uncomfortable to wear a mask because of their constantly runny nose, 88 (31%) because of cough, and 61 (22%) because of difficulty in breathing. 116 (40%) reported that their cough worsened when they wore a mask for a long period of time. Consequently, 197 (70%) reported sometimes taking off their mask to blow their nose or to cough. One in four (65 people) found communication difficult with people who wear a facemask. This was particularly pronounced among participants with hearing impairment due to PCD (46%). Most participants paid for masks themselves. 49 (17%) received facemasks from their workplace or from medical staff. Very few (4%) reported that they cannot afford as many masks as needed, but 51 (22%) reported that the cost of masks was burdening their budget.

**Table 3:** Beliefs about facemask effectiveness and problems related to wearing a face mask reported by people with PCD (N=282)

|  | Total |
| --- | --- |
|  | n (%) |
| **I disagree that:** |  |
| -masks protect me from getting infected with COVID-19 | 27 (10) |
| -masks protect me from spreading COVID-19 to others in case I am sick | 11 (4) |
| **I agree that:** |  |
| -when I wear a mask for a long time, my cough worsens | 116 (41) |
| **I find it uncomfortable to wear a mask because of the following:** |  |
| Runny nose | 94 (33) |
| Cough | 88 (31) |
| Difficulty breathing | 61 (22) |
| Headache | 7 (2) |
| Other problems<sup>a</sup> | 34 (12) |
| Any of the above (runny nose, cough, difficulty breathing, headache, other) | 137 (50) |
| **I must sometimes take my mask off because of runny nose or cough** | 197 (70) |
| **How frequently do I take my mask off because of runny nose or cough? (n=195)** |  |
| Every few minutes | 5 (3) |
| A few times per hour | 75 (38) |
| Rarely, at long intervals | 115 (59) |
| **I have problems communicating with others when they wear a mask** |  |
| Among all participants | 65 (23) |
| Among participants **with** hearing impairment (n=114) | 52 (46) |
| Among participants **without** hearing impairment (n=168) | 13 (8) |
| **Sometimes I am the only person who wears a mask in places where it is not mandatory** | 118 (42) |
| **Who pays for my masks (n=251)** |  |
| They are provided or reimbursed | 21 (8) |
| I pay them from my own pocket | 193 (78) |
| I pay them but I also get some provided or reimbursed | 28 (11) |
| I make them myself | 6 (2) |
| **Can I afford my masks (n=238)** |  |
| I cannot afford as many as I need | 8 (4) |
| Masks are heavy on my budget | 51 (22) |
| Masks fit in my budget | 176 (75) |

<sup>a</sup>Other problems included difficulty concentrating (n=2), sinus pain due to pressure from mask (n=2), glasses fogging (n=18), sweating (n=2), fitting problems due to hearing aids or problems with size (n=8), itchiness (n=1), fear of stigmatizing (n=1).

The only factor that was associated with not wearing a facemask in public was national facemask requirements (**table 4**). 17 (6%) of the participants reported that they never wore a mask in two or more public places and 12 of these were from countries where facemasks were not required anywhere or were required only in certain regions. Demographic factors, beliefs about effectiveness, problems related to mask wearing, self-reported lung function, or affordability were not associated with never wearing a facemask in public. Even participants who reported that people with PCD in their region were exempt from wearing a mask mostly reported to always wear a mask (91%). Of the two people who reported having a personal facemask exemption, one reported sometimes wearing a mask, and the other reported never wearing a mask.

**Table 4:** Factors associated with not wearing a mask in public among people with PCD (N=282)

|  | Wore a mask in public# | Never wore a mask in one public place | Never wore a mask in two or more public places | P-value* |
| --- | --- | --- | --- | --- |
|  | N=252 | N=13 | N=17 |  |
| **Age group** |  |  |  | 0.360 |
| 0-17 years | 64 (89) | 6 (8) | 2 (3) |  |
| 18-49 years | 132 (89) | 5 (3) | 11 (7) |  |
| Above 49 years | 56 (90) | 2 (3) | 4 (6) |  |
| **Sex** |  |  |  | 0.132 |
| Male | 87 (85) | 8 (8) | 7 (7) |  |
| Female | 164 (92) | 5 (3) | 10 (6) |  |
| **Exempt from wearing a mask because of PCD** |  |  |  | 0.004 |
| No | 161 (90) | 10 (6) | 7 (4) |  |
| Yes, personal exemption | 0 | 1 (50) | 1 (50) |  |
| Yes, people with PCD in my region are exempt | 29 (91) | 1 (3) | 2 (6) |  |
| I don't know | 61 (91) | 0 | 6 (9) |  |
| **Beliefs about wearing a mask (disagree or strongly disagree)** |  |  |  |  |
| Masks protect me from getting infected with COVID-19 | 23 (85) | 1 (4) | 3 (11) | 0.395 |
| Masks protect me from spreading COVID-19 to others | 8 (73) | 1 (10) | 2 (18) | 0.100 |
| When I wear a mask, my cough worsens | 104 (90) | 5 (4) | 7 (6) | 1.000 |
| **Uncomfortable to wear a mask because of cough, runny nose, or other reason (n=274)** |  |  |  | 0.780 |
| No | 124 (91) | 7 (5) | 6 (4) |  |
| Yes | 124 (91) | 5 (4) | 8 (6) |  |
| **Self-reported FEV<sub>1</sub> at study entry (n=192)** |  |  |  | 1.000 |
| FEV<sub>1</sub><sup>#</sup> above 40% predicted | 151 (87) | 11 (6) | 12 (7) |  |
| FEV<sub>1</sub><sup>#</sup> below 40% predicted | 16 (89) | 1 (6) | 1 (6) |  |
| **Difficult to communicate with others when they wear a mask** |  |  |  | 0.214 |
| No | 194 (89) | 12 (6) | 11 (5) |  |
| Yes | 58 (89) | 1 (2) | 6 (9) |  |
| **Affordability of masks** |  |  |  | 0.727 |
| I cannot afford as many as I need | 9 (100) | 0 | 0 |  |
| Masks are heavy on my budget | 44 (86) | 4 (8) | 3 (6) |  |
| Masks fit in my budget | 163 (92) | 8 (4) | 7 (4) |  |
| **Country specific facemask recommendation** |  |  |  | <0.001 |
| Facemasks required in whole country | 227 (93) | 12 (5) | 5 (2) |  |
| Facemasks required in parts of country | 20 (77) | 0 | 6 (23) |  |
| Facemasks not required | 5 (42) | 1 (8) | 6 (50) |  |

#Busy street, public transport, Bus-/train station grocery store, clothes store, restaurants, cinemas, hairdressers, physiotherapists, fitness studios, bank-/post offices \*Chi-squared test or Fischer's exact if cell-counts were less than 10.
#FEV<sub>1</sub>: forced exhaled volume in one second

## Discussion

This international participatory study found that people with PCD shielded themselves carefully against an infection with SARS-CoV-2. They avoided going to many public places and almost always wore facemasks in public. The public place which was visited most often by people with PCD was grocery stores where the majority always wore a mask (94%). Half of the participants experienced problems with wearing a face mask due to their chronic symptoms like recurrent cough, runny nose, and difficulty breathing, and a quarter of the participants reported that wearing a facemask worsened their cough. Only 2 people (1%) had a personal medical facemask exemption, and 32 people lived in regions with a general facemask exemption for persons with PCD. Nevertheless, most of them wore a facemask in public and did not take advantage of the exemption. Facemask wearing in public was not associated with negative beliefs about facemask effectiveness or problems experienced by wearing them, only by national facemask requirements.

A major strength of this study is the large sample size of people with a rare respiratory disease from different countries. It is difficult to recruit people with rare diseases for research, but the COVID-PCD is a participatory project that was initiated, designed, and tested in collaboration with people who have PCD. This boosted the study participation. Another strength is that the study is translated into five different languages (English, German, Spanish, Italian, and French). Our questionnaire included detailed questions about facemask use that made it possible to comprehensively understand facemask usage and related problems in a high-risk population. Our results are based on self-reported data, not real-life observations, but there was little risk of recall bias as the survey was sent out during a period when facemask use was compulsory in most countries and participants were wearing masks every day (October 2020). The study was anonymous so the risk of social desirability bias where answers would be influenced on what is socially accepted (e.g. all should wear masks) was low. Another limitation of our study was that only half of the population completed the questionnaire about mask use. Many participants were too young to wear a mask regularly and some parents may not have completed the questionnaire for this reason. This was confirmed by those who completed the facemask questionnaire being older than those who did not (**additional file 2**).

To our knowledge, this study is the first to describe facemask usage and related problems in people with a chronic disease during the COVID-19 pandemic. Few studies have reported on facemask usage in the general population, and they included only local populations making them unsuitable for direct comparison with our study [11, 28]. In a US survey of 1056 adults conducted in May 2020, 825 (79%) reported to wear a mask. The study included no information on places where facemasks were worn or frequency of use [28]. Similarly, in a cross-sectional online survey from Brazil conducted in July 2020, 1266 of 1277 (99%) said that they used facemasks but with no specification of where and how often [11]. The study further showed that two thirds of the participants (67%) were bothered by facemask in some way; 55% because of shortness of breath, 50% because of pain around the ears, and 44% because of glasses fogging. In our study, only 50% of people with PCD reported that facemasks were uncomfortable for any reason. A possible explanation may be that people with PCD are used to breathing problems in general and their threshold for what is uncomfortable may be higher. It may also be, that they are very conscious of the gains related to facemask use and thus more tolerant towards discomfort.

Studies show that risk perception, disbelief in face mask effectiveness, and presence of national facemask policies are associated with adherence to wearing facemasks [29, 30]. People with PCD were considered at high risk of severe COVID-19 and most of the participants in our study supported the opinion that facemasks are effective in preventing transmission of SARS-CoV-2. This may explain why almost all wore a facemask in public even if many reported problems because of chronic cough and rhinitis. The only factor that was associated with wearing a facemask in our study was lack of public policies on facemask requirement. Only two people had a personal medical exemption from wearing a facemask, and only one of them never wore a facemask in public. One in ten reported that people with PCD were exempt from wearing a face mask in their region, but most of these (91%) always wore a facemask in public despite this exemption. Among this group of people with a rare disease, of which many experienced breathing problems while wearing a mask, few exerted the right to not wear a mask in public. Facemasks are an important measure to reduce transmission of SARS-CoV-2 and, and facemask exemptions should be given based on individual needs and sparingly.

## Conclusion

This international study found that people with PCD carefully shield themselves, and most wear facemasks everywhere in public. People who did not wear facemasks in public came from countries without a national facemask requirement. National policies mandating facemask use in public are important for universal use to protect high-risk populations from SARS-CoV-2 infections.

## Data Availability

The COVID-PCD data is available on reasonable request by contacting Claudia Kuehni by

## Abbreviations

COVID-19: coronavirus disease 2019
PCD: primary ciliary dyskinesia
SD: standard deviation

## Declarations

### Ethics approval and consent to participate

The Bern Cantonal Ethics Committee (Kantonale Ethikkomission Bern) has approved this study (Study ID: 2020-00830). Informed consent to participate in the study is provided at registration into the study. Study participation is anonymous. Participants can withdraw their consent to participate at any time by contacting the study team.

### Availability of data and materials

The COVID-PCD data is available on reasonable request by contacting Claudia Kuehni by

### Authors’ contributions

ESL Pedersen, ERN Collaud, R Mozun, M Goutaki, and CE Kuehni made substantial contributions to the study concept and design. ESL Pedersen analysed the data and drafted the manuscript. ESL Pedersen, ERN Collaud, R Mozun, K Dexter, C Kruljac, H Silberschmidt, JS, Lucas, M Goutaki, and CE Kuehni critically revised and approved the facemask questionnaire and the manuscript.

## Acknowledgements

We thank all participants and their families, and we thank the PCD support groups and physicians who have advertised the study. We thank our collaborators who helped set up the COVID-PCD study: Cristina Ardura, Yin Ting Lam, Christina Mallet, Helena Koppe, Dominique Rubi, University of Bern. We thank Amanda Harris, University Hospital Southampton, for her contribution to the facemask questionnaire. We thank Francesca Santamaria, Federica Annunziata, and Vincenzo Miranda, Federico ll University, Naples, Italy, for helping to translate the study questionnaires to Italian.

---

**Additional file 1:** Formulation of questions and answers from the special questionnaire on facemask use sent to participants in October 2020.

| Topic | Question | Answer category |
| --- | --- | --- |
| Date | Date of completing the questionnaire | Date |
| Facemask wearing | *Does your child sometimes wear a face mask? | 1)No, my child is not old enough to wear a face mask<br>2)Yes, my child sometimes wear a face mask |
| Exemption | Are you exempt from wearing a mask because you have primary ciliary dyskinesia? | 1)No, I have to wear a mask where it is mandatory 2)Yes, I have a personal mask exemption prescribed by my doctor 3)Yes, people with PCD in my region can choose if they want to wear a mask 4)I don't know |
| Place and frequency of facemask wearing | Where and how often do you wear a mask?<br>Asked for the following public places: Grocery stores, Clothes shops, Restaurants, Bars, Public transport, Train stations, Cinemas, Hairdressers, Physiotherapy, Fitness centres, Yoga classes, Team sports, Contact sports, Bank-/post offices, My school, My workplace, Busy streets, Parks | 1)Always 2)Sometimes 3)Never 4)I don't go there |
| Kind of facemask | What kind of mask do you wear? | 1)Single-use mask certified 2)Single-use mask non-certified 3)Filtering face piece (FFP or FFP2/FFP3) 4)Fabric mask with exchangeable filters 5)Fabric mask without exchangeable filters 6)Other |
| Problems with facemasks | Do you find it uncomfortable to wear a mask because of your health problems? | 1)No, I do not find it uncomfortable 2)Yes, because of my runny nose 3)Yes, because of my cough 4)Yes, because of other health problems |
| Problems with facemasks | Are there other reasons that make it uncomfortable for you to wear a mask | Free text |
| Consequence of facemask | What do you usually do when you have a runny nose or cough and wear a mask? | 1)When I have a runny nose or cough, I avoid wearing a mask 2)I sniff in and swallow the mucus or suppress the cough and do not take the mask off 3)I sniff in and swallow the mucus or suppress the cough but I also take the mask off from time to time to clear my nose 4)I take the mask off frequently to clear my nose |
| Take off facemasks | How often do you take the mask off to clear your nose or spit out mucus when you have cough or a runny nose? | 1)Every few minutes 2)A few times per hour 3)Rarely, a long intervals |
| Beliefs about facemasks | How much do you agree with the following sentences?<br>-Masks protect me from getting infected by COVID-19<br>-Masks protect me from spreading COVID-19 to others in case I am sick<br>-When I wear a mask for a long time, my cough worsens because I suppress my cough or because I do not clear my nose | 1)I strongly agree 2)I somewhat agree 3)I neither agree nor disagree 4)I somewhat disagree 5)I strongly disagree 6)I don't know |
| Hearing | Do you have a hearing impairment? | 1)No 2)Yes |
| Communication | Do you find communicating with other people more difficult when they wear a mask? | 1)No 2)Yes, but I can deal with this problem quite easily 3)Yes, this is difficult for me |
| Only one wearing facemask | Are you sometimes the only one wearing a mask when it is not mandatory? | 1)No, other people also wear a mask in most places I go to 2) Yes, sometimes 3)Yes, often |
| Financing facemask | Do you have to pay for masks out of your own pocket? | 1)No, they are provided or I am reimbursed 2)Yes, I pay for them from my own pocket 3)Yes, mainly from my own pocket but some are also provided or I am reimbursed 4)Yes, but I make them myself |
| Affordability | Are masks affordable for you? | 1)No, I cannot afford to buy as many masks as I need 2) Yes, but they are heavy on my budget 3) Yes, they fit in my budget |
| Comments | Please write any additional comments you may have about mask use | Free text |

The questions described in this table were adapted to three age-groups: children aged 0-14 years (completed by parents), adolescents aged 14-17 years, and adults aged 18 or above. \*Only asked in the questionnaire for children aged 0-14 years

**Additional file 2:**
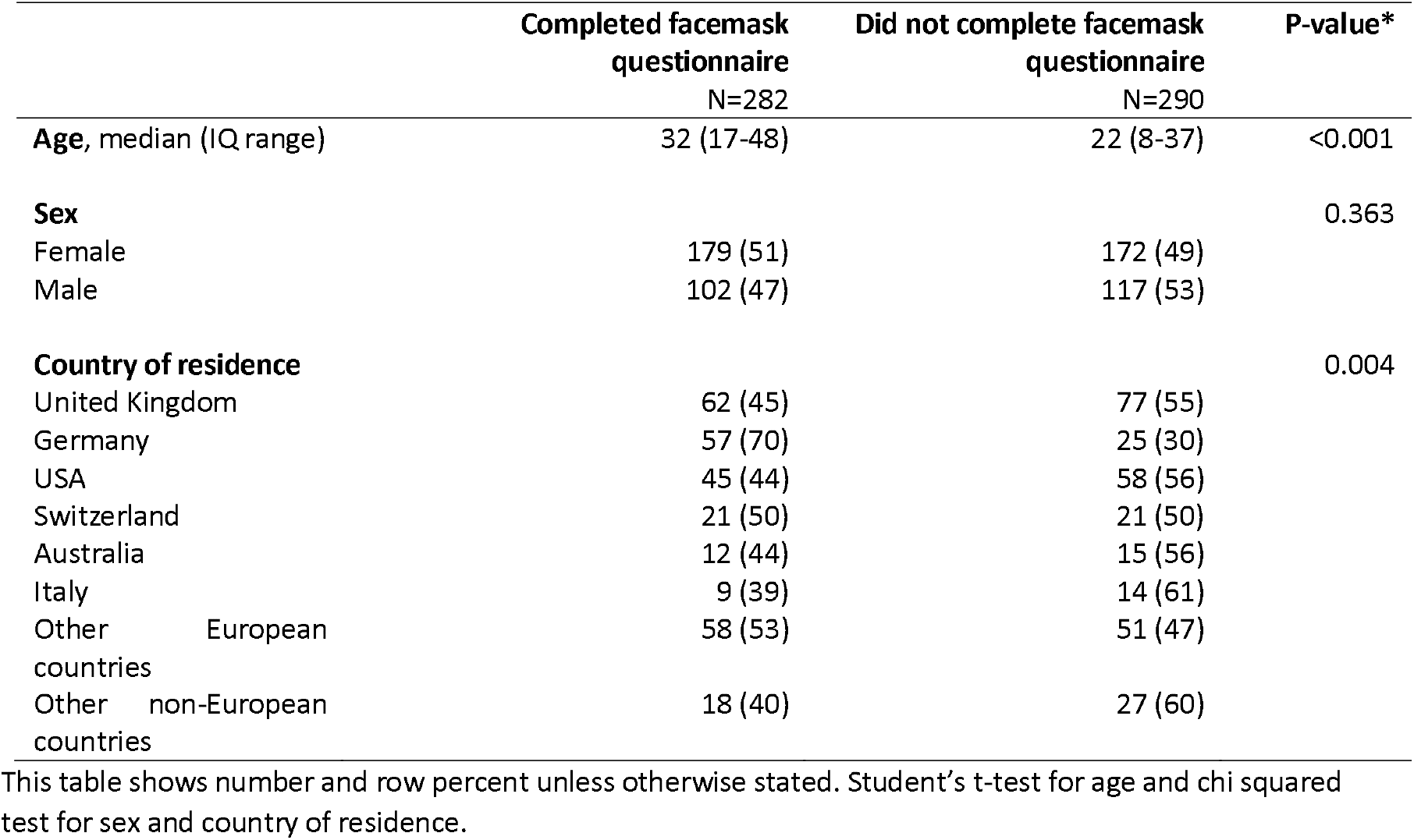
Comparison of people who completed the facemask questionnaire and those who did not among participants included in the COVID-PCD study by October 10, 2020 (N=572).

|  | Completed facemask<br>questionnaire<br>N=282 | Did not complete facemask<br>questionnaire<br>N=290 | P-value* |
| --- | --- | --- | --- |
| **Age, median (IQ range)** | 32 (17-48) | 22 (8-37) | <0.001 |
| **Sex** |  |  | 0.363 |
| Female | 179 (51) | 172 (49) |  |
| Male | 102 (47) | 117 (53) |  |
| **Country of residence** |  |  | 0.004 |
| United Kingdom | 62 (45) | 77 (55) |  |
| Germany | 57 (70) | 25 (30) |  |
| USA | 45 (44) | 58 (56) |  |
| Switzerland | 21 (50) | 21 (50) |  |
| Australia | 12 (44) | 15 (56) |  |
| Italy | 9 (39) | 14 (61) |  |
| Other European<br>countries | 58 (53) | 51 (47) |  |
| Other non-European<br>countries | 18 (40) | 27 (60) |  |

This table shows number and row percent unless otherwise stated. Student's t-test for age and chi squared test for sex and country of residence.

